# DEEP LEARNING TO PREDICT DEGREE OF INTERSTITIAL FIBROSIS AND TUBULAR ATROPHY FROM KIDNEY ULTRASOUND IMAGES – AN ARTIFICIAL INTELLIGENCE APPROACH

**DOI:** 10.1101/2020.08.17.20176958

**Authors:** Ambarish M. Athavale, Peter D. Hart, Mathew Itteera, David Cimbaluk, Tushar Patel, Anas Alabka, George Dunea, Jose Arruda, Ashok Singh, Avi Rosenberg, Hemant Kulkarni

## Abstract

**Background:** Interstitial fibrosis and tubular atrophy (IFTA) is a strong predictor of decline in kidney function. Non-invasive test to assess IFTA is not available.

**Methods:** We trained, validated and tested a deep learning (DL) system to classify IFTA grade from 6,135 ultrasound images obtained from 352 patients who underwent kidney biopsy. Of 6,135 ultrasound images, 5,523 were used for training (n = 5,122) and validation (n = 401) and 612 to test the accuracy of the DL system. IFTA grade scored by nephropathologist on trichrome stained kidney biopsy slide was used as reference standard.

**Results:** There were 159 patients (2,701 ultrasound images), 74 patients (1,239 ultrasound images), 41 patients (701 ultrasound images) and 78 patients (1,494 ultrasound images) with IFTA grades 1, 2, 3 and 4, respectively. The deep-learning classification system used masked images based on a 91% accurate kidney segmentation routine. The performance matrices for the deep learning classifier algorithm in the validation set showed excellent precision (90%), recall (76%), accuracy (84%) and F1-score (80%). In the independent test set also, performance matrices showed excellent precision (90%), recall (80%), accuracy (87%) and F1-score of (84%). Accuracy was highest for IFTA grade 1 (98%) and IFTA grade 4 (82%).

**Conclusion:** A DL system can accurately predict IFTA from kidney ultrasound image.

## Introduction

Interstitial fibrosis and tubular atrophy (IFTA) corelates with estimated glomerular filtration rate (eGFR)^1^ and the degree of IFTA is predictive of future decline in kidney function and development of end stage kidney disease^2^. Studies have shown that IFTA adds to predictive value of baseline proteinuria and eGFR in predicting outcome in patients with kidney disease^3^. IFTA predicts prognosis irrespective of underlying etiology of kidney disease^3,4^. Currently, histopathological assessment of kidney core biopsy by a pathologist is the only method to quantify IFTA that is routinely available in clinical practice. However, kidney biopsy is an invasive procedure, is associated with bleeding complications, provides a snapshot rather than continuous assessment of IFTA and is subject to sampling error^5,6^. Most importantly, the majority of chronic kidney disease patients who will progress to need renal replacement therapy (due to diabetes and hypertension)^7^ never undergo a kidney biopsy.^8,9^

A non-invasive method to quantify IFTA is not available, hence, unless a kidney biopsy is performed, the degree of IFTA is not available in for most patients. Kidney ultrasound is a routinely performed, non-invasive test in evaluation of kidney disease. Certain features on ultrasound such as echogenicity, kidney length and corticomedullary differentiation have been shown to corelate with IFTA; however, as currently performed and reported these features are unable to provide a quantifiable estimate of the degree of IFTA^10^. In contrast with FibroScan® which provides a quantification of fibrosis in liver^11^, there is no imaging modality that can provide an accurate estimate of IFTA in the kidney in routine clinical practice.

We hypothesized that encoded within ultrasonographic features are subtle IFTA correlates that can be quantitatively extracted and analyzed to derive an IFTA predictor. In this regard, it is noteworthy that artificial intelligence and deep learning have been utilized in diagnosis and prognosis of various medical conditions such as pulmonary tuberculosis from chest radiographs papilledema from fundus photograph and diagnosis of COVID pneumonia from CT scan^12–14^. Since deep learning has the ability to map complex feature relationships, we investigated whether a deep learning system could predict IFTA from ultrasound images of the kidney. Here, we trained, validated and tested a deep learning system to predict IFTA from kidney ultrasound images.

## Methods

### Patient selection

Consecutive patients undergoing native kidney biopsy between January 1, 2014 till December 31, 2018 were included in the study. Allograft biopsies were excluded. A total of 352 patients were included in this study. Clinical and demographic information of patients included in this study was obtained by chart review.

The study was approved by institutional review board at Cook County Health, Chicago, IL, USA.

### Quantification of Interstitial fibrosis and tubular atrophy

The percentage of cortex with interstitial fibrosis/tubular atrophy was estimated on Masson’s trichrome-stained sections of the renal cortex, and recorded for each biopsy. Quantifying IFTA on trichrome stained kidney biopsy slide is the current standard-of-care^15^. The percentage of cortex with interstitial fibrosis/tubular atrophy was scored as 0–24%, 25–49%, 50–74% and 74–100% of the cortex sampled. Two experienced renal pathologists (D.C., T.P.) independently performed the histologic evaluation.

Nephropathologist (DC) provided IFTA scores from each trichrome-stained histopathological slide of the kidney biopsy core. To validate the methodology of pathologist (DC) in grading IFTA, a second nephropathologist (TP) also provided IFTA scores for a random sample (N=93) of trichrome stained whole slide images in a blinded fashion and agreement between the two nephropathologists was tested. Clinical and demographic information of patients included in the study was obtained by chart review.

### Kidney Ultrasound images

Ultrasound studies of eligible patients were reviewed and all longitudinal images from both kidneys obtained within 6 months before and 2 weeks after kidney biopsy were included in the study. All kidney biopsies were performed under real time ultrasound guidance and ultrasound images obtained during the kidney biopsy were also utilized for the deep learning analysis. Ultrasound images were deidentified and stored in JPEG format.

### Development of deep learning classification system

Development of deep learning model involved four independent steps (Figure 1): 1. Preprocessing of ultrasound images, 2. Kidney segmentation, 3. Extracting features from segmented ultrasound images of the kidney, and 4. Image classification with internal and external validation. All scripts were written in Python 3.7 within Anaconda 3.0 environment^16^ and with PyTorch library^17^ as the backend. Jupyter Notebooks with codes and outputs are available from the authors upon reasonable request. Below, we describe each component in detail.

**Figure 1.**
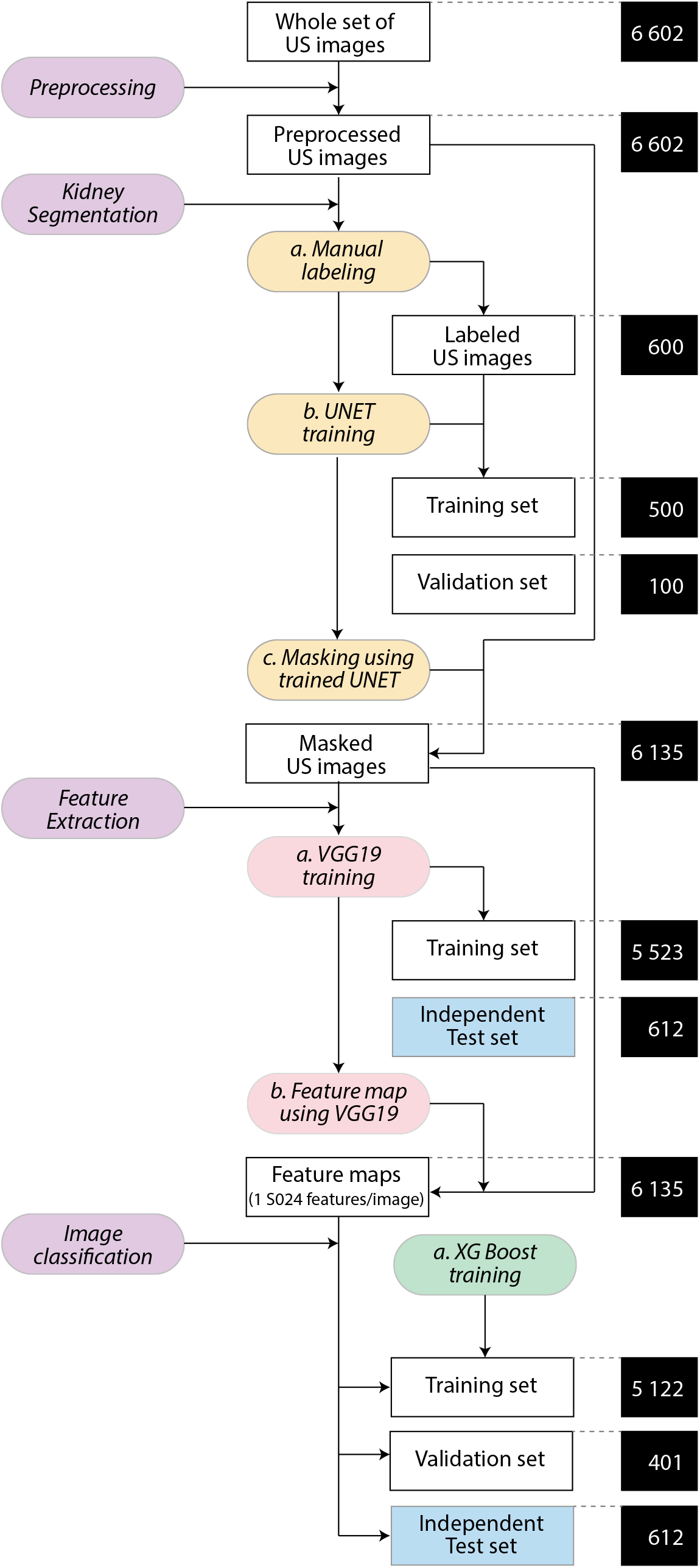
Overall analysis pipeline. The entire process was partitioned into four main tasks (purple colored boxes)- preprocessing of images, segmentation of kidneys in preprocessed images, feature extraction from masked images and image classification from feature maps. Sub-tasks within these main tasks are indicated with color-coded boxes. Unfilled rectangular boxes indicate the data inputs and black colored boxes (connected to the data input boxes by dashed lines) indicate the number of images used at that stage of the process. In the feature extraction and image classification phase a test set of 612 images (shown in the blue box) was generated and was never used in any training. This test set was used for a final independent evaluation of the overall analytical pipeline.

### Preprocessing of ultrasound images

Ultrasound images included in the study were resized to 224×224 pixel, an input dimension required for many popular pre-trained models. Crimmins filter^18^ (also called the geometric filter), is most suited to reduce background noise and backscatter in ultrasound images and was applied to each image.

### Kidney segmentation

A kidney ultrasound image may include other organs and surrounding structures in addition to the kidney such as muscle, adipose tissue, liver, spleen and bowel. For the purpose of this study, it was important that the training images contained only the kidney while eliminating other structures. For this, we first trained and validated a deep learning model to generate segmented (masked) ultrasound images. For this, we chose the UNet architecture (so called because of a series of downsampling followed by upsampling convolutions, Figure 3A) since it is suited for ultrasound images^19–22^. We used a pretrained model available publicly (https://github.com/jakeoung/Unet_pytorch) and retrained it for kidney segmentation. We randomly selected a subset of 600 ultrasound images (Figure 1) and manually labeled these images for kidney identification using the labelme^23^ software. The selected ultrasound images were further randomly split into a training set (n = 500) and a validation set (N = 100). Using this optimized (for mean squared error L2 loss) and trained UNet model we generated masked images (i.e. images with everything other than kidneys blacked out) from each of the 6602 preprocessed images. We used the intersection-over-union (IoU) metric to measure the accuracy of segmentation. We used the OpenCV Python library^24^ and used the function ‘multiply’ to obtain a masked image from the original image and its UNet-generated mask. Of 6602 images, 6135 images had adequate masks (IoU > 90%) and these masked images (n = 6,135) were used for subsequent analyses.

**Figure 3.**
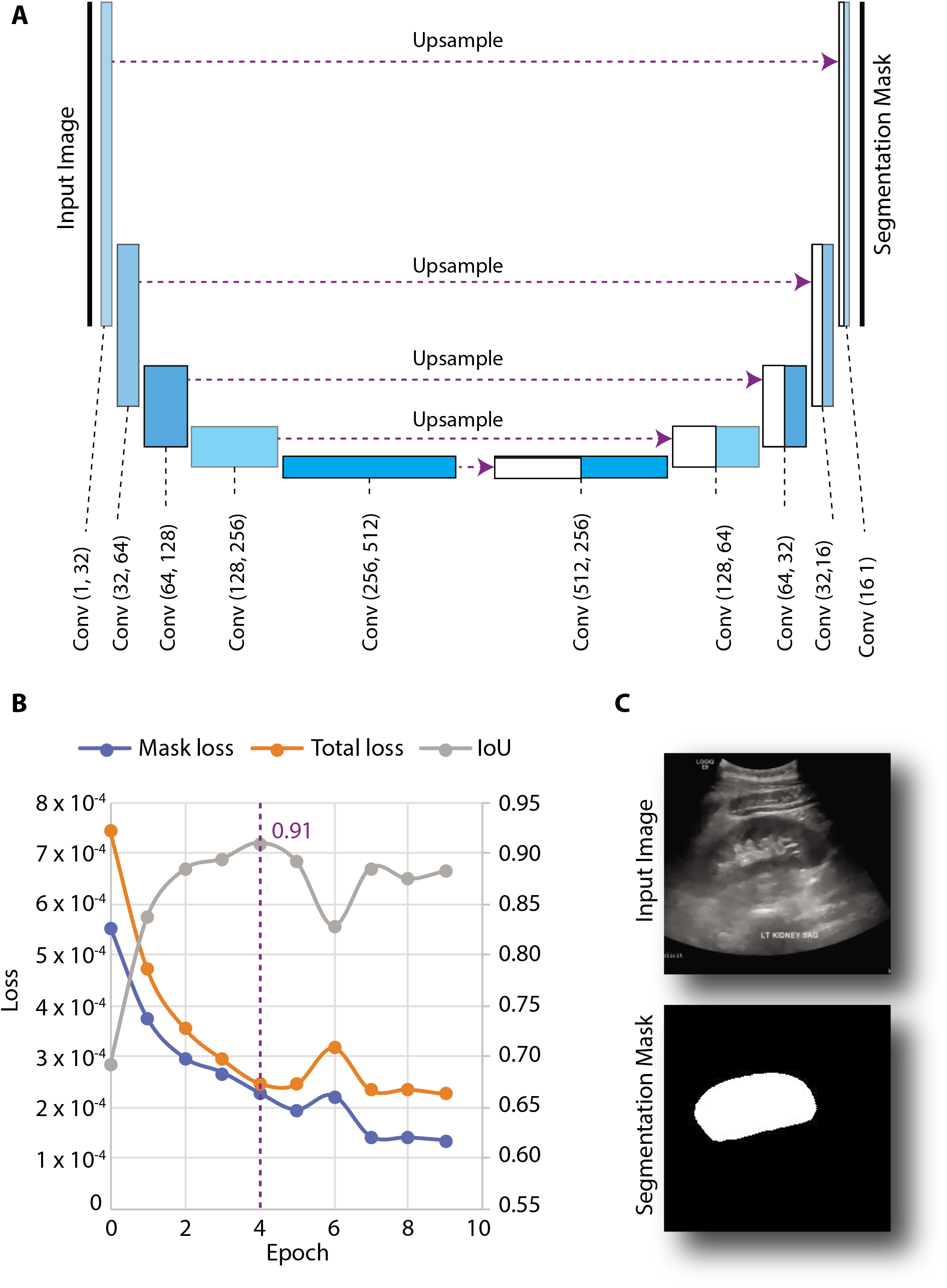
Use of UNet to predict kidney segmentation masks. (A) UNet architecture used. For details please visit: https://github.com/jakeoung/Unet pvtorch. The blue boxes represent convolution blocks with indicated number of input and output dimensions. Dashed purple arrows indicate upsampling paths. Input image (size 224 × 224 pixels) and generated masks are indicated by black vertical lines at the ends of the Unet architecture. (B) Plot of the two loss functions (mask-specific and total) and the loU (intersection over union) metric per training epoch. Maximum average loU (0.91) was observed after 4 epochs. (C) Example of an input and output generated using the trained Unet model.

### Feature extraction

We employed transfer learning for this purpose using a pre-trained convolutional neural network - VGG-19 BN (VGG - Visual Geometry Group, BN – batch normalization). This model (Figure 4A) comprises of an initial feature extractor component followed by a classifier component. For our purposes, we only used the feature extractor component. To prime it for kidney ultrasound fibrosis, we used histopathological grades as the final output and tuned the VGG-19 BN model using categorical cross-entropy cost function. From the trained model, we extracted 1,024 features (from the Fc2 layer) using the IntermediateLayerGetter Python library (https://github.com/sebamenabar/Pytorch-lntermediateLaverGetter). The final output of the 7×7×512 features was then flattened into a vector of 25,088 features and further compressed into a 1,024-length feature vector as shown in Figure 4A. Training of the feature extractor was done on a randomly selected subset of 90% images (N = 5,523, Figure 1). The remaining 612 images were retained as an independent test set for external validation. After training, the tuned VGG-19 BN model was used to extract features from all images into a 6,135 × 1,024 matrix. This matrix, along with the associated class labels, image identifiers and training/test membership information was used for subsequent image classification.

**Figure 4.**
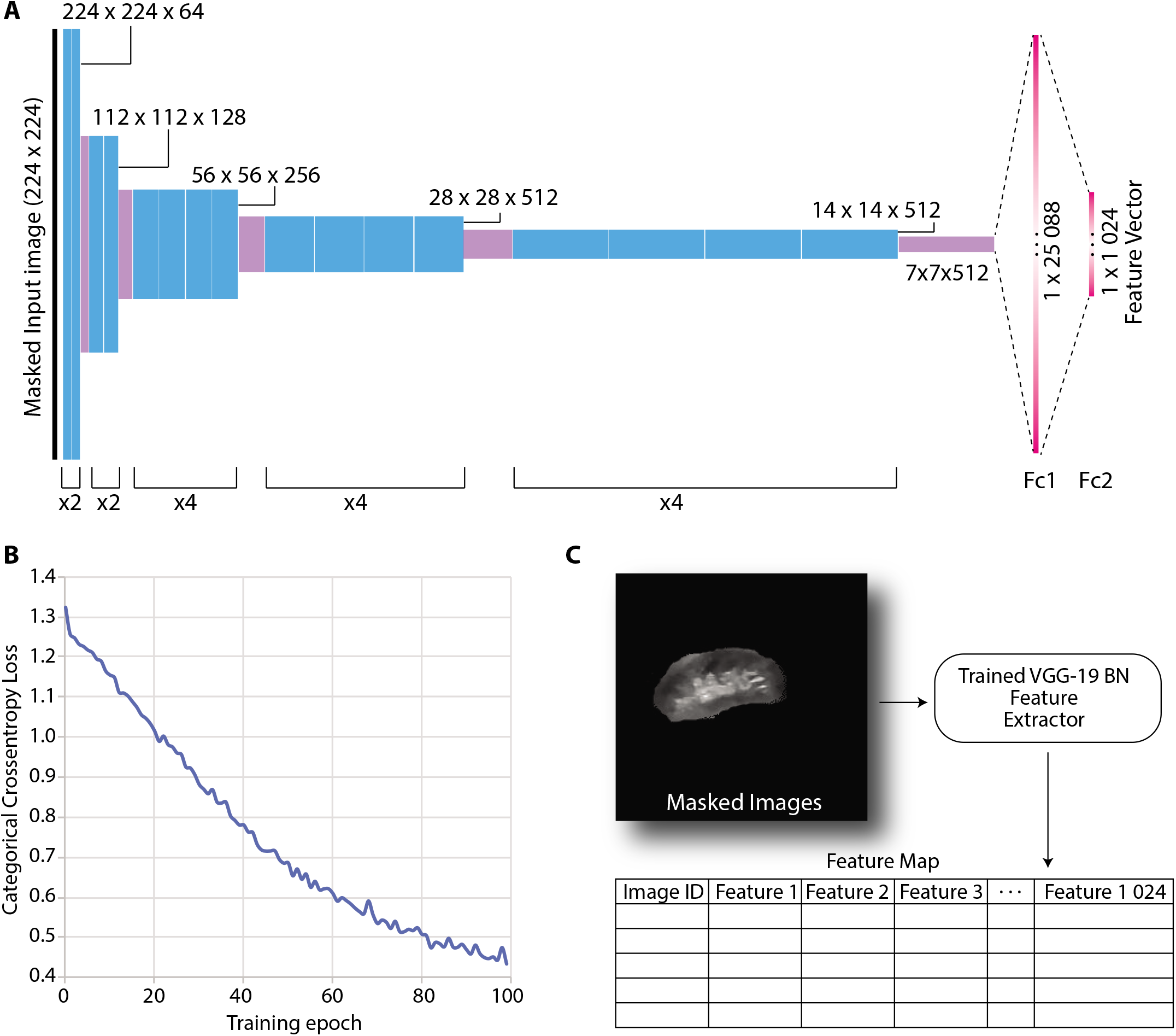
Use of VGG19-BN model for feature extraction. (A) The feature extractor portion of the VGG19-BN model. Input image is shown as black vertical line. Convolution operations are shown as blue boxes the width of which is proportional to the number of filters used for convolutions. Max pooling layers that reduced the size of the input image by half are shown as purple boxes. Fully connected layers (Fc1 and Fc2) were added for feature extraction. Number of convolution operations in each black are shown at the bottom. The output of this model was a 1,024-length feature vector (output of the Fc2 layer) for each image, contingent upon a final class assignment to 4 classes. (B) Plot of the categorical cross-entropy loos function during training for the first 100 epochs. (C) Use of the trained model. The trained VGG19-BN feature extractor was then applied to all the images (training as well as independent test sets) to generate a matrix containing the 1,024 features per image. This matrix was used for image classification in the subsequent step.

### Image classification

Image classification was done using extreme gradient boosting (XGBoosting using the xgboost Python library, https://github.com/dmlc/xgboost). During this step, we retained the 612 images as an independent test set (blue box, Figure 1). The remaining images were randomly split into a training set of 5,122 images and a validation set of 401 images. We used the 1024 features extracted in the feature extraction step as input to XGBoost algorithm and the ground truth histopathological grades as output for training the DL algorithm. Multiclass log loss was used for optimization. Grid search was used for finding the optimum hyperparameters and the best predicting model (one with the least multiclass error) was used to predict the IFTA grades both in the validation set (n = 401) and in the independent test set (n = 612).

### Statistical analysis

Descriptive statistics included mean and standard deviation for continuous variables and proportions for categorical variables. Statistical significance for distribution across grades of IFTA was assessed using one-way analysis of variance (ANOVA) for continuous variables and Pearson’s c2 for categorical variables. Agreement between pathologists’ grading of IFTA was done using weighted (using w^2^ strategy) Cohen’s kappa. Performance metrics for the image classification task were precision (synonymous with positive predictive value as used in epidemiology), recall (synonymous with sensitivity), accuracy and F1 score (which was estimated as the harmonic mean of precision and recall). All statistical analyses were conducted in Stata 12.0 (Stata Corp, College Station, TX) software package. A global type I error rate of 0.05 was used to test statistical significance.

## Results

### Study participants and ultrasound images

A total of 367 kidney biopsies were performed between January 1, 2014 to December 31, 2018 of which information on degree of IFTA as well as concurrent ultrasound images were available for 352 biopsies. Clinical and demographic characteristics of these patients are shown in Table 1. Number of patients assigned to different IFTA grades was as follows: grade 1, 159 (45.17%); grade 2, 74 (21.02%); grade 3, 41 (11.65%); and grade 4, 78 (22.16%). Consistent with clinical expectation, IFTA grade increased with age, presence of diabetes and hypertension and serum creatinine. IFTA grade was not associated with sex or race. For the 352 biopsies included in the study, a total of 6,135 ultrasound images had adequate “masks” (Figure 1) and were used to train and test the deep learning algorithm.

**Table 1.** Characteristics of the study participants

| Characteristic | IFTA<br><25%<br>N=159 | IFTA<br>25%-49%<br>N=74 | IFTA<br>50%-74%<br>N=41 | IFTA<br>≥75%<br>N=78 | p |
| --- | --- | --- | --- | --- | --- |
| Age (y) | 42.6 (13.4) | 52.9 (13.5) | 51.7 (13.7) | 49.8 (14.5) | <0.0001 |
| Females (%) | 93 (59.5) | 43 (57.1) | 21 (51.2) | 36 (48.2) | 0.356 |
| Race (%) |  |  |  |  | 0.155 |
| White | 70 (44.2) | 21 (31.2) | 10 (24.4) | 29 (38.6) |  |
| AA | 56 (35.6) | 39 (50.7) | 21 (51.2) | 37 (47.0) |  |
| Asian | 11 (6.8) | 7 (9.1) | 2 (4.9) | 5 (6.0) |  |
| Other | 22 (13.5) | 7 (9.1) | 8 (19.5) | 7 (8.4) |  |
| Diabetes | 39 (23.9) | 32 (44.1) | 18 (43.9) | 40 (50.6) | <0.0001 |
| Hypertension | 101 (63.8) | 68 (90.9) | 40 (97.6) | 71 (91.6) | <0.0001 |
| BMI (Kg/m<sup>2</sup>) | 29.7 (7.0) | 29.8 (7.0) | 29.2 (7.1) | 29.8 (6.3) | 0.8789 |
| Creatinine (mg/dl) | 1.49 (1.84) | 2.21 (1.43) | 2.41 (0.85) | 4.17 (2.18) | <0.0001 |
| eGFR (ml/min) | 87.3 (51.0) | 41.0 (24.3) | 30.8 (11.2) | 19.2 (10.1) | <0.0001 |
| Proteinuria (gm/gm creatinine) | 4.06 (3.9) | 4.92 (5.23) | 3.65 (2.76) | 4.90 (4.76) | 0.2330 |
| Biopsy diagnosis |  |  |  |  |  |
| Lupus nephritis | 62 (39.0) | 8 (10.8) | 3 (7.3) | 8 (10.3) | <0.0001 |
| Diabetic nephropathy | 4 (2.5) | 19 (25.7) | 10 (24.4) | 24 (30.8) | <0.0001 |
| FSGS | 14 (8.8) | 20 (27.0) | 12 (29.3) | 8 (10.3) | 0.0001 |
| IgA nephropathy | 19 (11.9) | 6 (8.1) | 4 (9.8) | 10 (12.8) | 0.7775 |
| Membranous GN | 21 (13.2) | 6 (8.1) | 2 (4.9) | 2 (2.6) | 0.0375 |
| ANCA vasculitis | 4 (2.5) | 4 (5.4) | 1 (2.4) | 9 (11.5) | 0.0237 |
| Hypertensive nephropathy | 1 (0.6) | 1 (1.4) | 2 (4.9) | 9 (11.5) | 0.0003 |
| Minimal change disease | 11 (6.9) | 2 (2.7) | 0 (0.0) | 0 (0.0) | 0.0242 |
| Number of ultrasound images |  |  |  |  |  |
| Total | 2,701 | 1,239 | 701 | 1,494 | 0.2012 |
| Per patient | 16.99 | 16.74 | 17.13 | 19.15 | 0.2849 |

IFTA, Interstitial fibrosis and tubular atrophy; y, years; AA, African American; BMI, Body mass index; eGFR, estimated glomerular filtration rate; FSGS, Focal segmental glomerulosclerosis; ANCA, Anti neutrophil cytoplasmic antibody

### Agreement between pathologists’ IFTA scores

Nephropathologist#1 (DC) provided IFTA grades for all 352 patients. Nephropathologist#2 (TP) independently provided IFTA grades for a random sample (N=93) of these 352 patients. Overall, there was excellent agreement between the two pathologists for IFTA classification. (Kappa of 0.84, p = 4.4×10–16, Table 2). However, the least frequent category of IFTA, grade 3 (50–74% IFTA score) was also the category with least agreement. Considering the overall excellent agreement between pathologists’ IFTA scores, we proceeded with the ensuing analyses using grades assigned by nephropathologist #1 (DC) (who had graded all the histopathology slides) as the ground truth labels for IFTA grades.

**Table 2.**
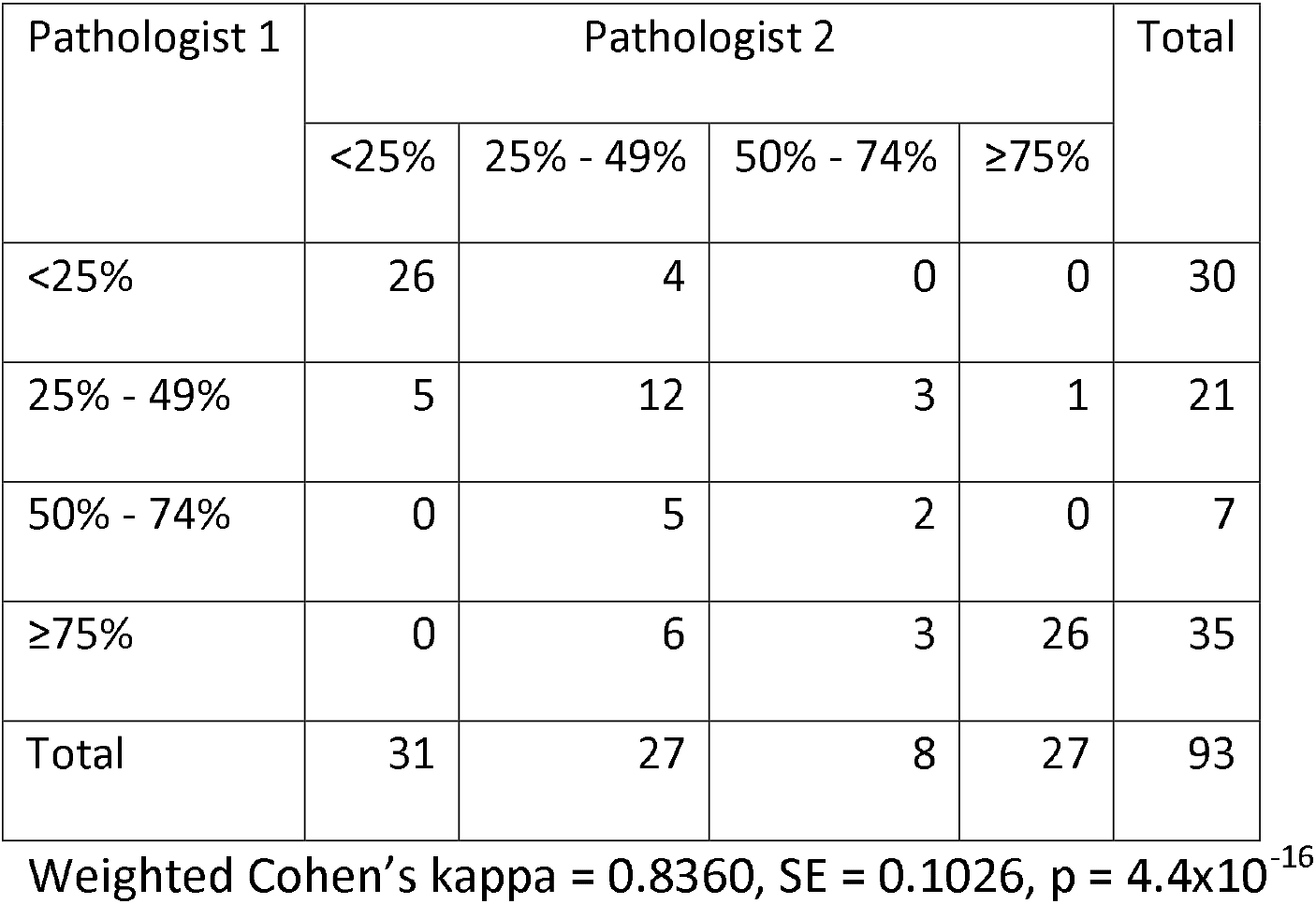
Agreement among pathologist’s independent evaluation of IFTA scores on randomly selected subsample of histopathology slides

| Pathologist 1 | Pathologist 2 |  |  |  | Total |
| --- | --- | --- | --- | --- | --- |
|  | <25% | 25% - 49% | 50% - 74% | ≥75% |  |
| <25% | 26 | 4 | 0 | 0 | 30 |
| 25% - 49% | 5 | 12 | 3 | 1 | 21 |
| 50% - 74% | 0 | 5 | 2 | 0 | 7 |
| ≥75% | 0 | 6 | 3 | 26 | 35 |
| Total | 31 | 27 | 8 | 27 | 93 |

Weighted Cohen's kappa = 0.8360, SE = 0.1026, p = $4.4 \times 10^{-16}$

### Preprocessing, kidney segmentation and feature extraction

We first preprocessed all the images with resizing and Crimmins filtering. As shown in Figure 2, the resulting images were smoother and of a standard size amenable to further processing. The next step of training a UNet model for kidney segmentation needed only 4 epochs to provide the best estimate of IoU with a rapidly decreasing loss (best IoU = 0.91, Figure 4B). We then subjected all the preprocessed images to this tuned Unet model. We inspected the resulting images and their masks (as shown in Figure 3C) manually and found that in poorly segmented images the proportion of the mask to the entire image was < 0.05. We thus excluded these images (n = 256) and retained a set of 6,346 that related to the entire set of 367 patients. After further excluding images from the 15 patients who did not fulfil the inclusion criteria, a set of 6,135 ultrasound images remained that were used for feature extraction using the VGG-19 BN pretrained model. Of these images, 5,523 were used for training the feature extractor. The training of the feature extractor was consistent, gradual and reasonably smooth (as shown by the decreasing loss function (Figure 4B). Using the tuned model, we generated the feature map for all the 6,135 masked images as shown in Figure 4C.

**Figure 2.**
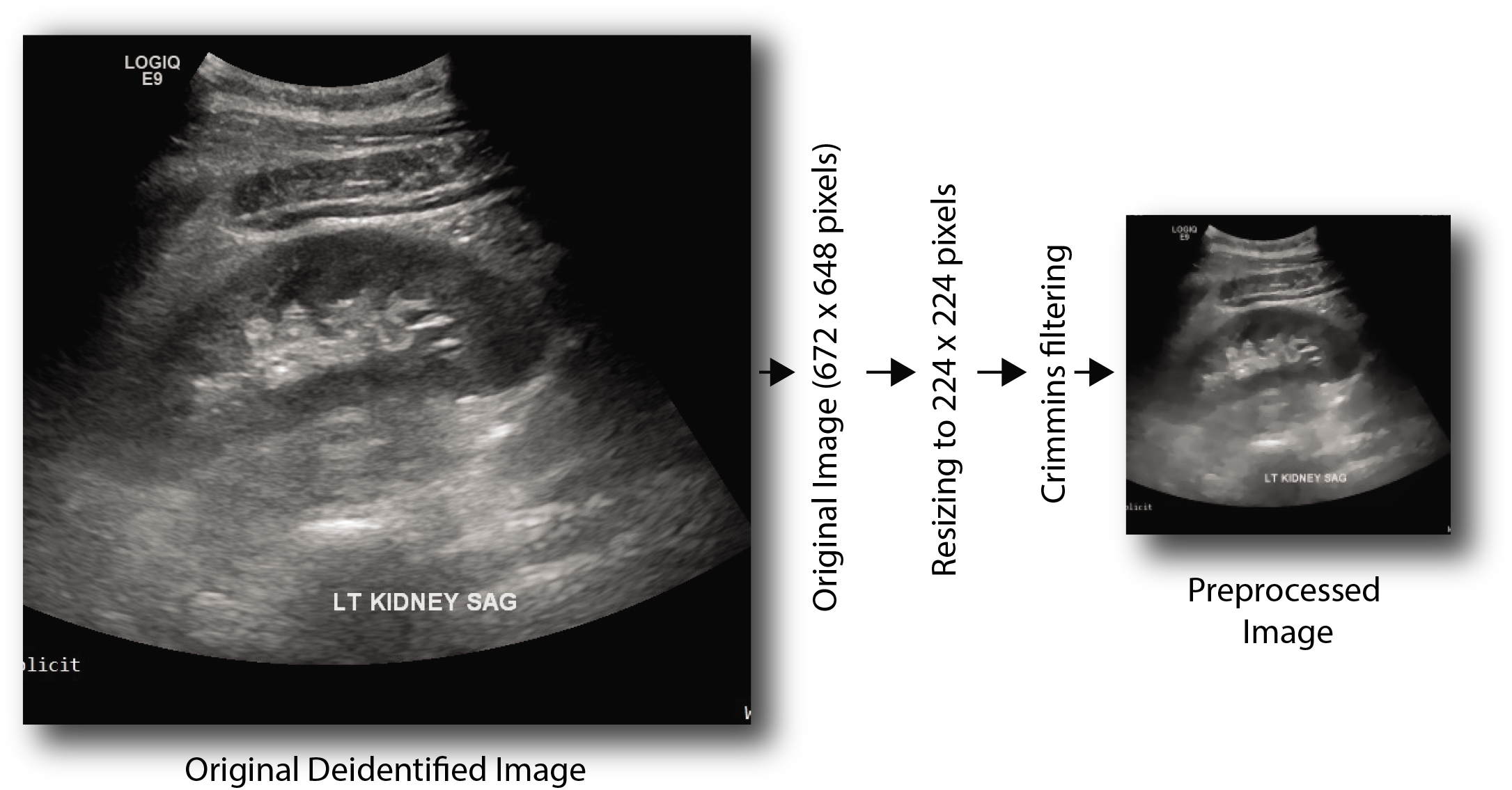
Typical preprocessing protocol. Deidentified images were first resized to 224 × 224 grayscale images and then subjected to Crimmins filtering. An example of input and preprocessed image is shown.

### Image classification

Using the feature map generated in the previous step as input, we then trained an XGBoost classifier. An exhaustive grid search yielded an optimal classification solution with the following set of hyperparameters: learning rate (eta) 0.01, maximum tree depth 16, subsample fraction 0.5 and a severe L2 regularization penalty (lambda) of 10. The decrement in loss function was monotonic and smooth in both training (n = 5,122) and validation (n = 401) sets as shown in Figure 5A. Concordantly the multiclass labeling accuracy consistently increased in both sets (Figure 5B) implying acceptable fit to the data. When this model was evaluated in the validation set (Figure 5C, we found that confusion matrix was dense along the diagonals and yielded a set of very good performance metrics (Figure 5D) with high precision (89.36%), good recall (76.46%), an accuracy of 84.29% and an F1-score of 80.54%. To further demonstrate the robustness of this approach, the image classifier was evaluated in the independent test set. We again observed a very similar performance in this test set (Figures 5E and 5F) with precision of 89.27%, good recall 80.37%, an accuracy of 86.75% and an F1-score of 83.89%. A closer look at the confusion matrices (Figures 5C and 5E) showed that the accuracy of prediction was highest for IFTA grade 1 (almost perfect) and IFTA grade 4 (81% in validation and 82% in test set) and lowest for the IFTA 3 grade (51% in validation and 66% in test set).

**Figure 5.**
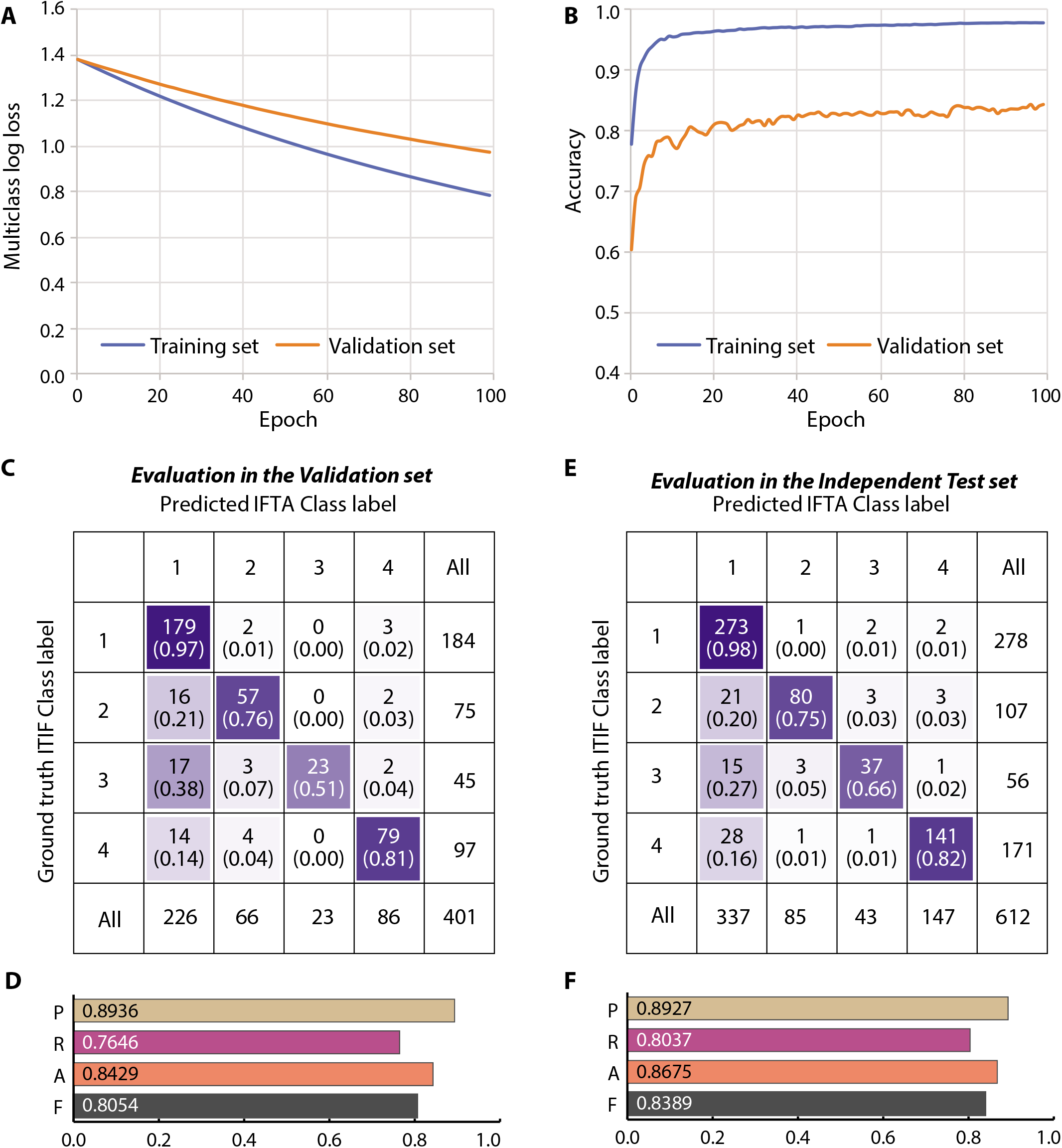
Results of image classification using XGBoost algorithm. Feature vectors generated in the previous step were used for training on 5,122 training images and 401 validation set (leaving aside the 612 images in the Independent Test set shown in blue box in Figure 1). (A) Multiclass log loss and (B) predictive accuracy in the training set (blue curves, n = 5,122) and validation set (orange curves, n = 401). Best prediction was observed for epoch 99. (C-D) Results of evaluation in the validation set (n = 401). Panel C shows the confusion matrix – numbers in each cell are count (proportion of the predicted labels within the corresponding actual labels). The cells are color coded to indicate the degree of accuracy – darker cells indicate stronger accuracy. Panel D shows bar plots for four studied metrics from the best classifier model – P, precision; R, recall; A, accuracy; and F, F1 score. Numbers inside the boxes show the estimated metrics. (E-F) Results of evaluation in the independent test set. The legends for panels E and F are similar to those for panels C and D, respectively.

## Discussion

We developed, validated and tested a convolutional neural network (Deep learning) based algorithm to predict IFTA (a histopathology-based classification) from ultrasound images of the kidney. To our knowledge, no such model currently exists although thinking in this direction using computerized tomography^25^ (CT) and magnetic resonance^26^ (MR) images have been made. Our prediction system capitalizes on ultrasound imaging which is done in patients with kidney disease irrespective of need for kidney biopsy. The overall diagnostic accuracy of the DL algorithm ranged from 84% in the validation set to 87% in the test set and the algorithm predicted the IFTA grade with very good accuracy when IFTA was < 25% and > 75%.

IFTA corelates with kidney function and is a strong predictor of future decline in kidney function and need for renal replacement therapy^2,3,27,28^. At present, information on IFTA grade is available only in patients who undergo kidney biopsy. However, kidney biopsy is an invasive procedure, can be associated with significant bleeding complications and is usually reserved for patients with proteinuria/hematuria/rapidly declining kidney function in whom a glomerular disease other than diabetes is suspected^5,6^. There have been prior attempts to corelate ultrasound findings with IFTA on kidney biopsy. O’Neal et al corelated ultrasonographic findings such as kidney length, echogenicity, parenchymal and cortical thickness with interstitial fibrosis and tubular atrophy findings on kidney biopsy^10^. They reported that kidney length, echogenicity and parenchymal thickness showed significant correlation with IFTA but the correlation was modest at best (0.35 for echogenicity and interstitial fibrosis). None of the ultrasonographic findings individually or in combination was able to provide a quantitative estimate of IFTA. Other ultrasonographic techniques such as quantitative echogenicity, shear wave velocity imaging, transient elastography and ultrasound corticomedullary strain have been evaluated^10^. However, unlike FibroScan which grades Liver fibrosis by transient elastography and has obviated the use of biopsy in chronic viral hepatitis, none of the current ultrasonographic methods can provide a clinically useful estimate of IFTA grade^29,30^. Several serum and urinary biomarkers have been evaluated as a noninvasive measure of IFTA but no biomarker has been sufficiently accurate to be useful in routine clinical practice. Thus, in a vast majority of patients with kidney disease, a non-invasive method to estimate degree of IFTA is needed. In our study, the deep learning algorithm was able to predict IFTA grade with 87% accuracy which is sufficient to be considered useful in clinical practice.

How would non-invasive prediction of IFTA be useful in clinical practice. To answer this question, it is relevant to look at utility of Liver Fibroscan as a corollary. Prior to advent of Fibroscan, Liver biopsy was needed to assess fibrosis and distinguish cirrhotic from non-cirrhotic Liver^11^. Fibroscan is able to non-invasively assess fibrosis and distinguish cirrhotic from non-cirrhotic Liver and Liver biopsy is no longer performed in most patients with Hepatitis B and C^31^. It is pertinent to note that the Fibroscan has good accuracy in detecting cirrhotic Liver but accuracy in detecting earlier stages of Liver fibrosis is moderate at best^31^. This is still helpful to the clinician in making treatment decisions for hepatitis B and C since presence or absence of fibrosis guides choice of therapy. The deep learning algorithm developed in our study is able to identify patients with IFTA < 25% or > 75% with high degree of accuracy. It is generally accepted that patients with advanced (> 75% fibrosis) are not good candidates for immunosuppressive therapy in proteinuric glomerular diseases. A non-invasive method of estimating fibrosis can potentially help in treatment decision without the need for invasive kidney biopsy. At present, there are more clinical trials evaluating treatment for proteinuric kidney diseases than at any time in the past. A non-invasive method to assess fibrosis like the one proposed in our study can potentially be helpful in selection of subjects for clinical trials.

Our study has significant strengths. We used a deep learning algorithm approach that was able to accurately detect complex non-linear features embedded within ultrasound images which is an advantage over conventional linear methods that attempt to corelate kidney length, echogenicity, parenchymal and cortical thickness with IFTA grades. Indeed, while conventional methods of reporting kidney ultrasound are sub-optimal in providing a quantification of IFTA grade, our method based on deep learning approach was able to quantify IFTA grade with overall accuracy of 87%. We used >6000 ultrasound images to train and validate the algorithm. The biopsy diagnosis represents the entire spectrum of kidney disease and no major category of kidney disease was excluded (other than cystic diseases of the kidney for which biopsy is typically not performed). However, our results should be considered in the light of some implicit limitations. This was a retrospective, observational study since the kidney biopsy and ultrasound studies were performed prior to this analysis. This has resulted in an imbalance in the proportion of patients in each IFTA grade. Although the overall diagnostic accuracy of 87% is high, the diagnostic accuracy in IFTA grade 3 was low. IFTA grade 3 also had the least number of subjects (11% of total sample) and also the least number of ultrasound images for training the algorithm (700) as compared to the other three IFTA grades. It is conceivable that the diagnostic accuracy for IFTA grade 3 would improve with a higher representation of ultrasound images in this grade. Interestingly, the weakest agreement between pathologists was for the IFTA grade 3 indicating the possibility of an inherent difficulty in assigning histopathological images to this grade. The pathologist who provided IFTA grades had access to the patient’s clinical history and laboratory findings but the deep learning algorithm utilized just the ultrasound images to provide an estimate of IFTA grade. It is possible that combining the clinical information with the ultrasound images in the deep learning algorithm will increase the accuracy of the IFTA grade estimate. The choices for available pretrained models are aplenty. Our choice of the VGG-19 BN model was driven by the motivation to use as simple models as possible but other deeper models like those belonging to the ResNet, DenseNet or Inception families may improve the accuracy of IFTA estimate. Our UNet segmentation model provided a high average IoU but it is likely that if this accuracy is further enhanced, it may lead to an improved feature extraction and classification ability of the system. Future studies are needed to improve the segmentation component of our system. Although the DL algorithm was validated on an independent sample of images, further validation on external datasets is needed. Finally, deep learning models are – by nature – adaptive. We anticipate a continually improved performance of the system as more and more real time data is provided for its continued learning.

In conclusion, we have developed an artificial intelligence-based and deep learning-driven algorithm that was trained on ultrasound images to predict IFTA grade with high degree of accuracy. Our paper provides proof of principle that an artificial intelligence deep learning system can be used to non-invasively predict IFTA grade in patients with kidney disease. While the system cannot be seen as an alternative to kidney biopsy, non-invasive assessment of IFTA has the potential to significantly enhance clinical decision making and prognostication in patients with kidney disease.

## Data Availability

Jupyter Notebooks with codes and outputs are available from the authors upon reasonable request.

## Author contributions

AMA and HK designed the study, drafted and revised the manuscript. HK conceptualized and designed the network architecture, conducted deep learning modeling and the statistical analysis. AMA, MI, AA gathered the data. DC, TP and AR provided IFTA grades from kidney biopsy slides and helped in critical revision of the manuscript. PH, JA, GD and AS critically revised the manuscript. All authors approve the final version of the manuscript.

## Acknowledgement

We acknowledge funding support from the Hektoen institute of Medicine, Chicago, IL for this study.

## Disclosures

The authors report no relevant conflict of interest.

## Notes

### Competing Interest Statement

The authors have declared no competing interest.

### Author Declarations

The study was approved by institutional review board at Cook County Health, Chicago, IL, USA.

